# Sex-Stratified Multi-Omics Identifies Sexually Dimorphic Molecular Targets in Parkinson’s Disease

**DOI:** 10.64898/2026.04.10.26350571

**Authors:** Joo-Yeon Lee, Jimin Lee, Seokho Lee, Jung Han Yoon, Don Gueu Park, Joohon Sung

**Author notes:** Correspondence to: Don Gueu Park: Ajou University School of Medicine, Suwon, Gyeonggi-do, 16499, South Korea, Joohon Sung: 220-704, Gwanak-ro 1, Gwanak-gu, Seoul, 08826, South Korea. These authors jointly supervised this work (Don Gueu Park, Joohon Sung).

## Abstract

Parkinson’s disease (PD) exhibits well-established sex differences in prevalence and clinical phenotypes, yet the underlying molecular mechanisms remain largely elusive. Here, we conducted a comprehensive sex-stratified multi-omic integration to identify sex-specific causal proteins and biological pathways in PD.

We performed gene-based association analysis, transcriptome-wide association studies (TWAS), and proteome-wide Mendelian randomization (PWMR) with colocalization analysis using GWAS summary statistics from the International PD Genetics Consortium (IPDGC; 12,054 male cases/11,999 controls; 7,384 female cases/12,389 controls) for sex-stratified analyses and Global Parkinson’s Genetics Program (GP2; 34,933 cases/31,009 controls) for sex-combined analyses. Prioritized candidates were further evaluated through MR with brain expression quantitative trait loci (eQTLs) from MetaBrain and differential protein abundance analysis using the Global Neurodegeneration Proteomics Consortium (GNPC; 704 PD cases/5,629 controls in plasma; 78 cases/1,411 controls in cerebrospinal fluid). Additionally, pathway enrichment analysis was performed for prioritized molecules.

Integration across three analytical layers prioritized 102 molecular candidates across 31 unique loci, significant from multiple analyses. Of these, eleven genes reached significance across all three layers, including *SNCA, MAPT,* and *CTSB* significant in both sexes; *CD160, GPNMB,* and *LRRC37A2* as male-predominant; *STX4* and *PRSS53* as female-predominant; and *BST1, SCARB2,* and *LGALS3* significant only in sex-combined analysis. In males, *CD160* emerged as a novel candidate with convergent evidence across all three analyses and colocalization, while *L3MBTL2* was identified as a novel risk gene from gene-based association and TWAS analyses. In females, *STX4* and *PRSS53* at the 16p11.2 locus showed female-predominant associations. Pathway enrichment analysis revealed innate immune and SUMOylation pathways in males, with *CD160* and *L3MBTL2* as key contributors respectively, contrasting with WDR5-mediated chromatin remodeling in females. Brain eQTL-based MR confirmed significant associations for 69 of 86 testable candidates (80.2%) in at least one tissue. Protein abundance analysis confirmed sex-specific patterns, and several candidates showed discordant directions between genetically predicted causal effects and observed protein abundance — including male-specific plasma elevation of *CD160* and female-specific patterns for *STX4* — underscoring the distinction between causal risk mechanisms and disease-state molecular changes.

These findings demonstrate that PD is a molecularly heterogeneous disorder with sexually dimorphic pathogenic drivers. While shared axes such as lysosomal dysfunction and vesicle trafficking disruption exist, the divergence into male-specific immune dysregulation and female-specific chromatin remodeling suggests that the primary triggers of neurodegeneration differ by sex. Our results underscore the necessity of sex-stratified approaches in biomarker discovery and the development of precision therapeutic strategies for PD.

## Introduction

Parkinson’s disease (PD) is the world’s fastest growing neurological disorder,^1^ affecting approximately 8.5 million individuals worldwide.^2^ Despite extensive global efforts to understand the pathophysiology of PD, its underlying mechanisms remain unknown, partly due to clinical heterogeneity. Accumulating evidence suggests that this heterogeneity is intimately linked to the patient’s sex.^3–7^

The sex differences in PD are evident in both epidemiological and clinical domains. Epidemiologically, PD prevalence is approximately 1.5 times higher in men.^8^ This finding is particularly notable as it contrasts with the pattern observed in other late-onset neurodegenerative disorders, such as Alzheimer’s disease, which exhibits a higher prevalence in women.^9^ This disparity suggests a relatively greater PD burden in men. Clinical presentations also differ by sex: male patients more frequently show rigidity-dominant motor phenotypes and faster cognitive decline, whereas female patients more often exhibit tremor-dominant disease, depression, and other non-motor symptoms.^5,6^

Research attempting to understand the mechanisms underlying these sex differences has focused on genetic/epigenetic factors, environmental exposure, and sex hormonal influences. Estrogen, in particular, is known to have a protective effect, supported by clinical observations such as accelerated PD onset in postmenopausal women and evidence suggesting hormone replacement therapy may reduce PD risk.^10,11^ Furthermore, the neuroprotective effect of the estrogen-related receptor gene expression associated with REM (rapid eye movement) sleep behavior disorder (RBD) and the potential contribution of testosterone to PD risk have also been investigated.^12^

Studies exploring underlying mechanisms of PD using omics data have revealed significant insights into its sexual dimorphism. The largest sex-stratified PD GWAS to date, conducted by the Global Parkinson’s Genetics Program (GP2), found that while the genetic architecture of PD is broadly similar between sexes, sex-specific analyses identified five novel risk loci.^13^ Beyond genomics, a sex-stratified Mendelian randomization (MR) study assessed the causal relationships between immune-related proteins and PD risk by sex, identifying several sex-specific proteins.^14^ Our recent drug-target MR study also identified the sex-specific effects of dipeptidyl peptidase-4 in male PD only.^15^ Several studies have investigated the differential gene expression and regulation with transcriptomics and epigenomic data.^16,17^

However, most prior sex-stratified studies of PD have been limited to a single molecular layer, leaving the convergence between genetic risk, gene expression, and protein levels in men and women largely unexplored. To address this gap, we integrated sex-stratified gene-based association, transcriptome-wide association, and proteome-wide Mendelian randomization analyses to dissect the sex-specific genetic architecture of PD and to nominate causal protein targets of potential therapeutic relevance.

## Materials and methods

### Data sources

We utilized GWAS summary statistics from two major PD consortia. For sex-combined analyses, we employed the latest GWAS results from the GP2.^18^ The GP2 GWAS is assembled from two sources — clinically-recruited case-control cohorts and population-based biobanks — and has been reported as a source-stratified meta-analysis. Leveraging this framework, for our three primary analyses we used the clinical-based cohort as the discovery dataset and the biobank-based cohort as an independent replication dataset. Subsequent downstream analyses were performed using the pooled dataset to maximize statistical power.

For sex-stratified analyses, as full summary statistics from recent GP2 GWAS^13^ were not publicly available at the time of analysis, we leveraged established data from the International Parkinson’s Disease Genetics Consortium (IPDGC). This dataset included 12,054 PD cases and 11,999 controls for men, and 7,384 cases and 12,389 controls for women.^19^ To minimize potential confounding by population stratification, all analyses were restricted to individuals of European ancestry (Supplementary Table 1).

### Gene-based association analysis

To identify genes associated with PD risk, we performed gene-based association analyses using MAGMA (v1.10).^20^ GWAS summary statistics were mapped to the human genome reference (GRCh37 for sex-stratified analysis and GRCh38 for combined analysis) to calculate gene-level p-values. Genome-wide significance was defined using Bonferroni-correction based on the number of genes.

We conducted both sex-combined and sex-stratified analyses. For the sex-combined dataset, signals identified in the discovery cohort were required to replicate at p-value < 0.01 in the replication cohort. To account for linkage disequilibrium (LD), we computed genetic distances using the 1000 Genomes Phase 1 European (CEU) reference panel,^21^ and considered loci independent when separated by more than 0.1 cM. Novelty of our findings was evaluated using Open Targets Genetics.^22^ Loci were classified as novel when the prior association score was < 0.2, and candidate genes were further prioritized using the locus-to-gene (L2G) score, which reflects predicted causal relevance.

### Transcriptome-wide association study (TWAS)

To identify genes associated with PD at the mRNA expression level, we performed a TWAS using FUSION software.^23^ By integrating PD GWAS summary statistics with expression quantitative trait loci (eQTL) reference weights derived from the GTEx (v8) dataset, we estimated the association between predicted gene expression and PD risk across 49 human tissues.^24^

Statistical significance was defined using tissue-specific Bonferroni correction (p < 0.05/number of tested genes per tissue). To ensure robustness and minimize tissue-specific noise, we prioritized cross-tissue consistent genes, defined as those reaching significance in at least two distinct tissues. Consistent with our gene-based association analyses, we conducted both sex-stratified and two-stage sex-combined analyses, further prioritizing identified genes using L2G and association scores from Open Targets Genetics.^22^

### Proteome-wide Mendelian randomization and colocalization analysis

To prioritize potentially causal proteins in PD pathogenesis, we conducted a PWMR analysis. Genetic instrumental variables (IVs) for plasma protein levels were constructed using *cis*-protein quantitative trait loci (pQTL) data from the UK Biobank Pharma Proteomics Project (UKB-PPP), in which protein levels were measured using the Olink platform.^25^ Independent SNPs (r^2^ < 0.1) within a ± 250 kb window of each gene boundary reaching suggestive significance (p < 1×10⁻^5^) were selected as IVs. SNP-exposure association coefficients from UKB-PPP were used as weights, and effect estimates are reported as odds ratios per unit increase in genetically predicted protein levels (rank-inverse normal transformed scale).

MR analysis was performed using the ‘TwoSampleMR’ (v0.6.22) R package,^26^ primarily using the inverse variance weighted (IVW) method (or Wald ratio when only a single SNP was available). PWMR analyses were performed for each sex separately, and for the sex-combined analysis using the two-stage framework described above. Proteins reaching false discovery rate (FDR) < 0.05 were retained for further evaluation. For these candidate proteins, sensitivity analyses were performed using complementary MR methods — MR-Egger regression, weighted median, and weighted mode — and MR with varying LD clumping thresholds (r^2^ < 0.05 and 0.01, in addition to 0.1) to assess the robustness of the causal estimates. Heterogeneity among instrumental variables was evaluated with Cochran’s Q test, and horizontal pleiotropy was assessed via the MR-Egger intercept. MR-PRESSO was additionally applied to detect potential outlier variants (outlier test p < 0.05), and MR analyses were repeated after excluding identified outliers.^27^ Instrument strength was assessed using the F-statistic, and F-statistic > 10 was considered to indicate sufficient instrument strength. Steiger directionality test was performed to verify that the genetic instruments explained more variance in the exposure (protein levels) than in the outcome (PD risk), confirming the correct causal direction. Proteins with a p-value < 0.01 were considered significant; for replication of the sex-combined analysis, a p-value < 0.05 with a concordant direction of effect was required.

For the causal proteins identified via MR, we performed colocalization analysis to assess whether genetic associations for protein level and PD risk share a common causal variant. Using the ‘coloc’ (v6.0) R package,^28^ we estimated posterior probabilities (PP) for five hypotheses (H_0_-H_4_) within a ± 250 kb window around the gene boundary. Evidence of colocalization was defined as a PP_H4_ (hypothesis for a shared causal variant) > 0.8. Prior probabilities implemented in the coloc R package were used (p1 = p2 = 1×10⁻^4^, p12 = 1×10⁻^5^). The MR analyses in this study were conducted and reported in accordance with the STROBE-MR guidelines.

### MR with eQTL from brain tissues

Using the candidates prioritized from the three primary analyses, we performed MR analyses with brain tissue-specific eQTLs. eQTL summary statistics were obtained from the MetaBrain consortium,^29^ which provides data from five brain regions (basal ganglia, cerebellum, cortex, hippocampus, and spinal cord). For each candidate gene, we selected independent *cis*-eQTLs (p < 10^−5^, r^2^ < 0.1, within ±250 kb of the gene boundary) as instrumental variables and conducted MR analyses following the same approach as described above.

### Differential protein abundance analysis

To validate our prioritized candidates at the translational level, we utilized proteomic profiles from the Global Neurodegeneration Proteomics Consortium (GNPC), which comprises over 35,000 biofluid samples.^30^ Proteins were quantified using the SomaScan platform in plasma or cerebrospinal fluid (CSF). We included six cohorts with at least 30 PD cases, encompassing a total of 704 PD cases (43.6% female) and 5,629 cognitively normal controls (59.1% female). Among these six cohorts, two cohorts also had CSF proteomic data, including 78 cases and 1,411 controls.

For each individual cohort, we performed linear regression adjusting for age and sex (for sex-combined analysis) or age only (for sex-stratified analysis). For the cohort without internal control data, pooled controls were utilized, consistent with the methodology of the original GNPC paper.^30^ Individual cohort results were then integrated via a weighted Z-score meta-analysis. To account for potential inter-cohort variability, we applied a fixed-effect model when heterogeneity was low (I^2^ < 50%) and a random-effects model when substantial heterogeneity was detected (I^2^ ≥ 50%).

### Pathway enrichment analysis

To interpret the biological implications of our multi-omic findings, we integrated results from the three primary axes (gene-based association, TWAS, and PWMR). We prioritized molecules significant in at least two of the three analyses. Sex-specific candidates were defined as those exhibiting significance in only one sex. Pathway enrichment analysis was subsequently performed using Reactome v95 (nominal p-value < 0.01).

## Data availability

This study utilized publicly available summary-level data. Sex-stratified PD GWAS summary statistics were obtained from the International Parkinson’s Disease Genetics Consortium (IPDGC; https://pdgenetics.org). Sex-combined PD GWAS summary statistics were obtained from the Global Parkinson’s Genetics Program (GP2) through the Neurodegenerative Disease Knowledge Portal (https://ndkp.hugeamp.org). Plasma cis-pQTL data were obtained from the UK Biobank Pharma Proteomics Project (UKB-PPP) available on the Synapse platform (https://www.synapse.org/Synapse:syn51365303), and brain eQTL data were obtained from MetaBrain (https://www.metabrain.nl/). TWAS reference weights were derived from GTEx v8 multi-tissue expression data by the Mancuso Lab, publicly available at http://gusevlab.org/projects/fusion/#gtex-v8-multi-tissue-expression. Proteomic data were obtained from the Global Neurodegeneration Proteomics Consortium (GNPC), available at the Alzheimer’s disease data initiative (https://www.alzheimersdata.org/).

## Results

The analytical framework and multi-omic integration process are illustrated in Figure 1. For the gene-based association analysis, we evaluated 19,014 genes in the sex-combined dataset and 18,308 genes for the sex-stratified analyses (Supplementary Table 2 and Supplementary Figure 1). In the combined analysis, we identified 146 genes across 44 independent loci that were successfully replicated. Notably, 17 loci were novel, having no prior association score or a score below 0.2 in Open Targets Genetics. Sex-stratified analyses identified 36 genes (12 loci) in males and 44 genes (9 loci) in females after Bonferroni correction. While 5 regions were shared between sexes, we identified several sex-specific signals; four loci (groups 1, 19, 46, and 48; Supplementary Table 2) were uniquely associated with male PD, while two loci (groups 29 and 33) were female-specific.

**Figure 1.**
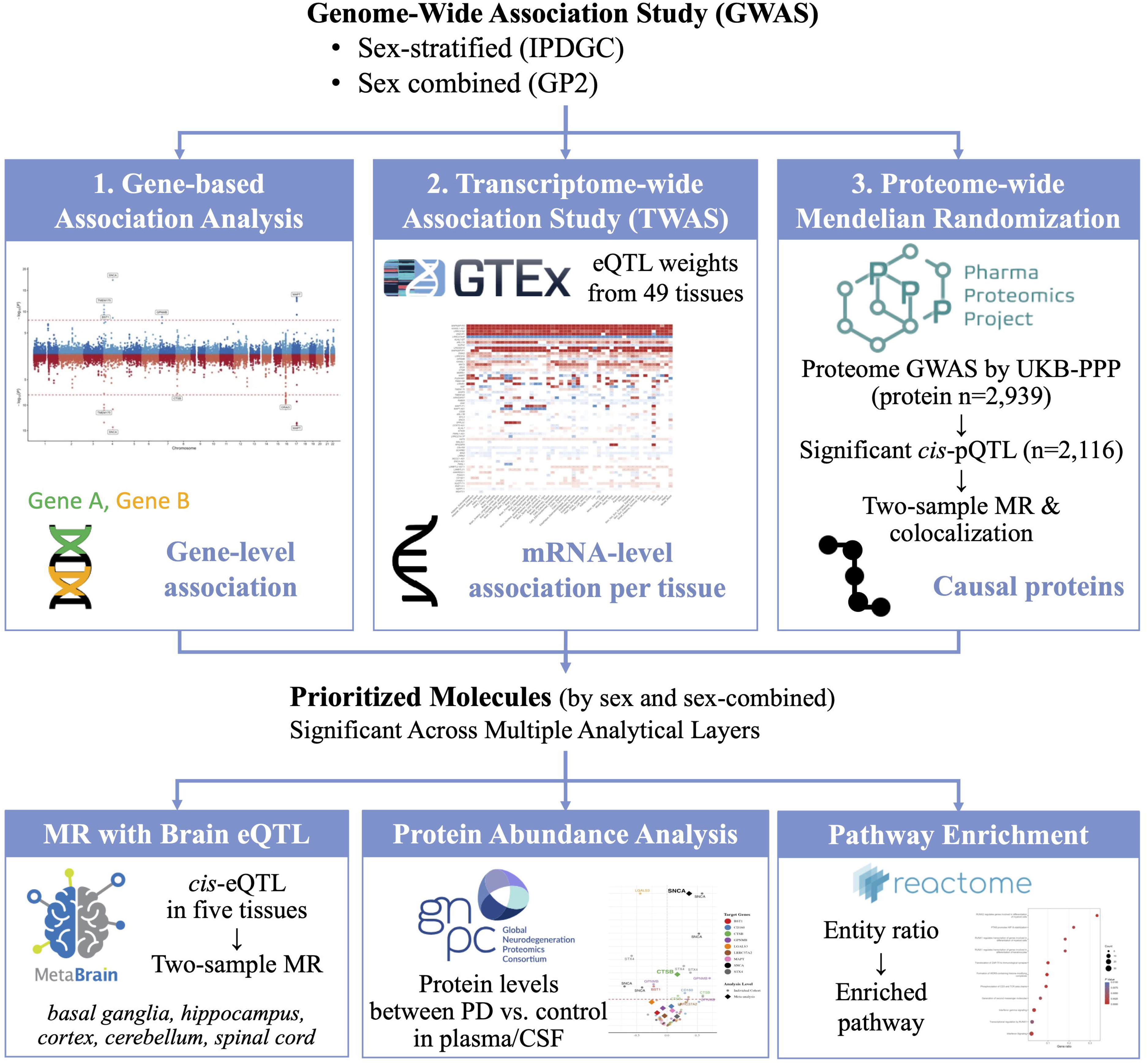
Integrative multi-omic framework for unraveling PD pathogenesis. This schematic illustrates the analytical pipeline for integrating genomic, transcriptomic, and proteomic data Three primary analyses were performed: (1) gene-based association analysis; (2) transcriptome-wide association study (TWAS); and (3) proteome-wide Mendelian randomization (PWMR) using *cis*-pQTLs, followed by colocalization analysis. Molecular candidates reaching significance across multiple analytical layers were prioritized and subjected to downstream analyses: MR with brain *cis*-eQTLs; differential protein abundance analysis in plasma and CSF; and pathway enrichment analysis.

We further expanded our investigation using TWAS to identify risk genes at the mRNA expression level. For significant genes from at least two tissues, their minimum p-value across 49 tissues for each gene was shown in Supplementary Table 3. In the sex-combined analysis, we identified 202 genes across 51 independent regions, including 25 novel loci (association score < 0.2). Sex-stratified TWAS identified 67 genes (17 loci) significantly associated with male PD and 73 genes (13 loci) with female PD. These findings included three male-specific regions (group 2, 36, and 54), among which groups 2 and 54 overlapped with the male-specific loci identified in the gene-based association analysis. Significant results from at least two tissues in sex-stratified TWAS across 49 tissues are shown in Supplementary Figure 2. Five genes showed consistently significant results in nearly all (> 45) tissues in both sexes: *KANSL1-AS1, MAPK8IP1P2, LRRC37A2,* and *DND1P1* were negatively associated with PD risk, whereas *LRRC37A4P* showed a positive association. In contrast, some genes showed inconsistent direction of effect across tissues, including *MAPT, PLEKHM1, CRHR1, ARHGAP27,* and *SNCA*.

Then we conducted PWMR analysis using proteome data (Supplementary Table 4). From the GWAS of 2,939 proteins in UKB-PPP, we could construct genetic instruments for 2,116 proteins (p-value < 10^−5^ and within 250 kb *cis*-region of each gene). In the sex-combined analysis, PWMR identified 65 significant proteins, of which 32 were replicated in an independent dataset. Sex-stratified analyses revealed 51 male-specific and 52 female-specific proteins, along with 25 proteins significant in both sexes and 15 significant only in the sex-combined analysis, yielding 143 candidate causal proteins in total. Among the 25 shared proteins, 24 exhibited consistent effect directions across sexes (15 protective, 9 deleterious). Subsequent colocalization analysis of these 143 proteins revealed that among 25 shared proteins, six proteins (*HLA-DRA, CTSB, HIP1R, CTF1, STX4,* and *PRSS53*) showed evidence supporting a shared causal variant (Supplementary Table 5). For sex-specific candidates, three male-specific proteins (*CD160, FCGR2A,* and *LRRC37A2*) and two female-specific proteins (*PON2* and *RSPO1*) showed strong evidence of colocalization.

For 17 genes consistently significant in PWMR and at least one other analytical layer, their MR results and sensitivity profiles are presented in Figure 2 and Supplementary Table 6. Although several instrumental variables showed evidence of heterogeneity or horizontal pleiotropy after excluding outliers, complementary MR methods robust to these violations — including weighted median/mode and MR-Egger — yielded consistent results. The exception was *GPNMB* in males, where evidence of pleiotropy was detected (MR-Egger intercept p = 0.026) and the pleiotropy-adjusted estimate was attenuated (IVW OR = 1.24, p = 4.8×10⁻^5^; MR-Egger OR = 1.13, p = 0.072). All instrumental variables demonstrated sufficient strength (F-statistic > 10), indicating minimal risk of weak instrument bias. Steiger directionality test confirmed the correct causal direction (protein → PD risk) for most candidate proteins, although directionality could not be statistically confirmed for proteins with limited instrument strength (e.g., those with a single instrumental variable) (Supplementary Table 6). Additionally, MR analyses using stricter clumping thresholds (r^2^ < 0.05 and 0.01) yielded consistent results, although several signals attenuated or disappeared for r^2^ < 0.01.

**Figure 2.**
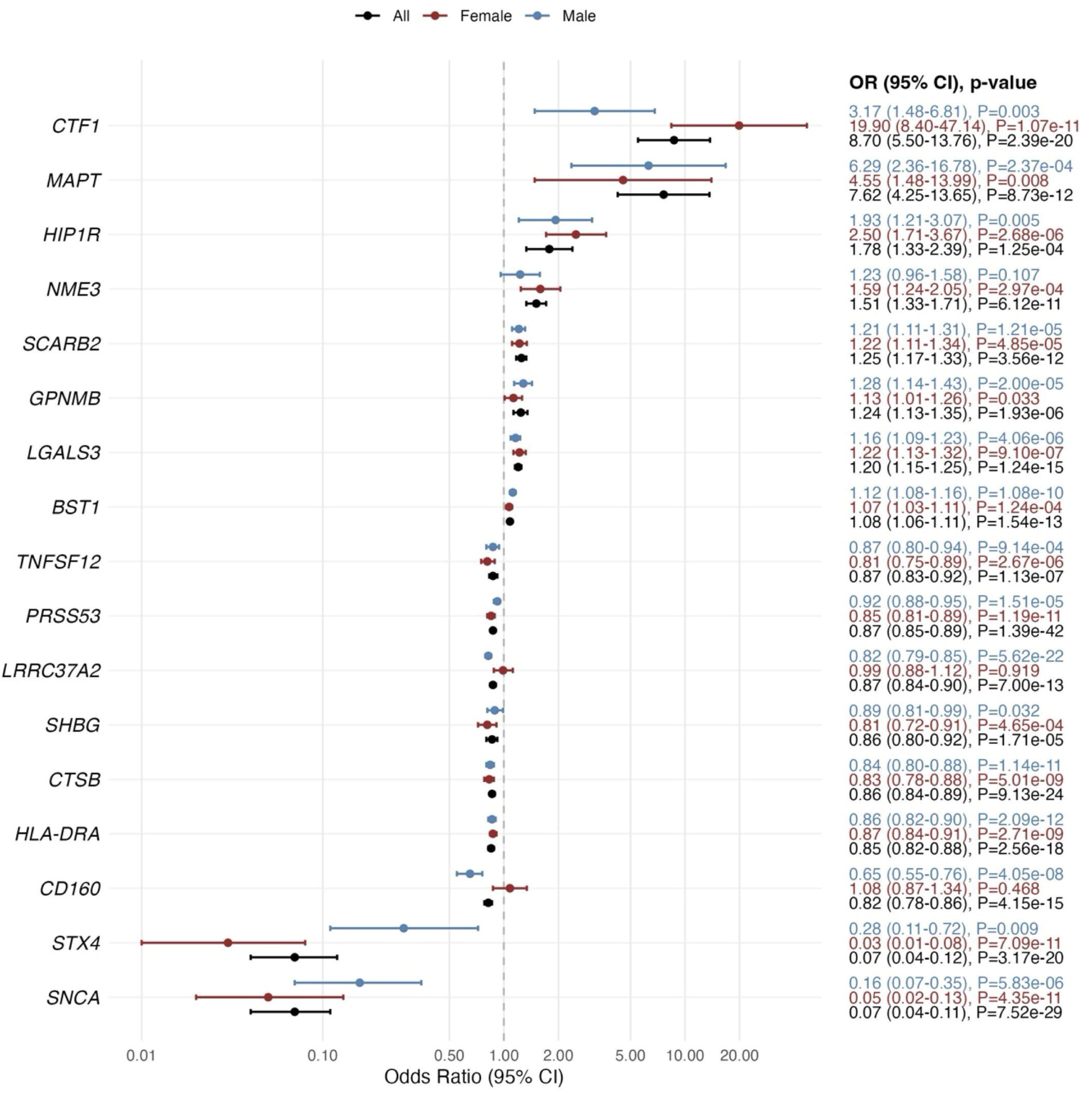
Proteome-wide Mendelian randomization (PWMR) identifying causal proteins for PD. Forest plots illustrate the causal effect sizes (Odds Ratio, OR) and 95% confidence intervals (CI) of candidate proteins on Parkinson’s disease (PD) risk. Proteins were selected as candidates based on a significant association in PWMR analysis (p < 0.01) across any of the analyzed strata (sex-combined, male, and female). To ensure the robustness of the identified causal signals, the inclusion was restricted to proteins whose encoding genes also demonstrated significant associations in either gene-based association analysis or TWAS.

By integrating results from the three primary multi-omic analyses, we prioritized 102 molecular candidates across 31 unique loci that reached significance in more than one analytical layer (Supplementary Table 8 and Figure 3). These comprised 28, 31, and 94 molecular genes spanning 10, 8, and 28 loci from male, female, and sex-combined analyses, respectively, after accounting for linkage disequilibrium. Three of the ten loci identified in male-stratified analyses were novel, and two loci (groups 1 and 10; Supplementary Table 8) were specifically associated with males, with p-values > 0.2 in female across all analyses. The genes with the highest L2G score in these two regions were *ANKRD35* and *L3MBTL2*. Although these specific genes were not represented in the PWMR analysis, *CD160* — located within the same locus as *ANKRD35* — exhibited a significant association with decreased PD risk specifically in males through PWMR (OR = 0.65, p = 4.1 × 10^−8^; Supplementary Table 4). In contrast, none of the eight female-associated loci were specific to women, although several genes such as *STX4* and *PRSS53* showed female dominance with suggestive associations in males.

**Figure 3.**
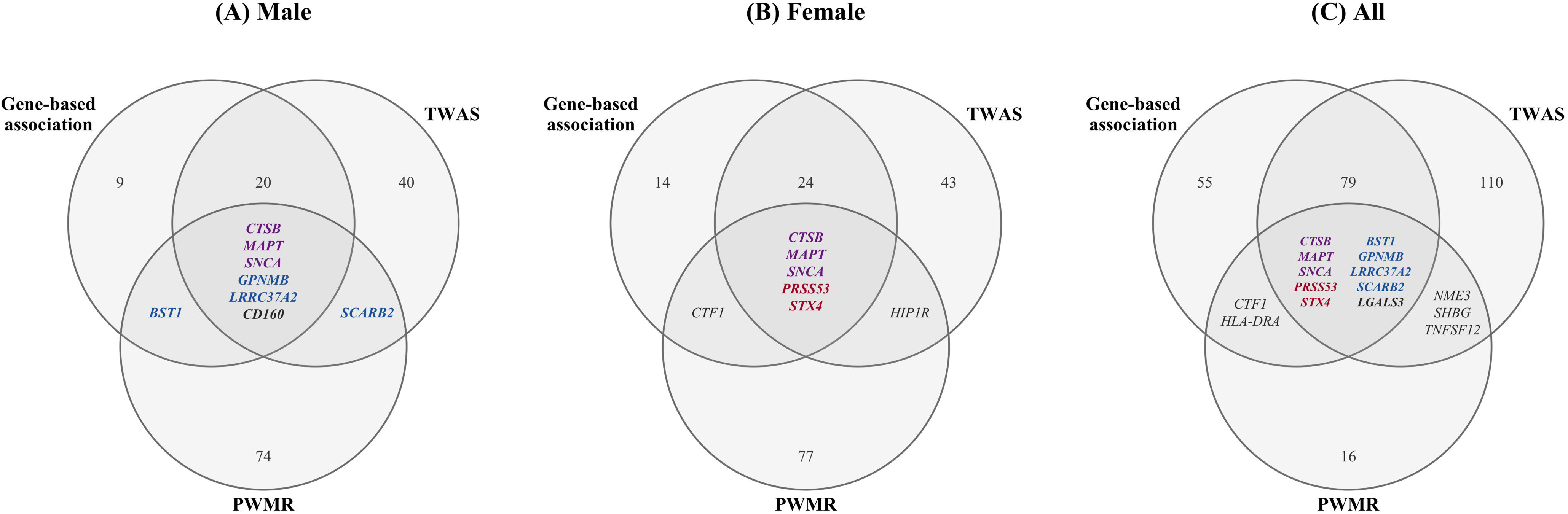
Intersection of PD-associated risk loci across genomic, transcriptomic, and proteomic layers. Venn diagrams illustrate the overlap of significant genes identified through gene-based association analysis, transcriptome-wide association study (TWAS), and proteome-wide Mendelian randomization (PWMR) in (A) male, (B) female, and (C) all PD. Numbers indicate the count of genes significant in each analysis or intersection. For intersections involving PWMR, individual gene names are annotated. Genes significant across all three analytical layers are shown in bold italic: purple denotes genes identified in both male and female analyses (*CTSB, MAPT,* and *SNCA*), blue denotes male-predominant genes (*BST1, GPNMB, SCARB2,* and *LRRC37A2*), and red denotes female-predominant genes (*PRSS53* and *STX4*).

Among the 102 candidates significant from multiple analyses, 86 had at least one significant *cis*-eQTL from five brain tissues (Supplementary Table 9). Of these, 69 (80.2%) showed significant associations with PD risk in at least one tissue and sex stratum. Notably, *SNCA,* which showed robust associations in both sexes using plasma pQTL instrument, had no significant results in either cerebellum or cortex, with data unavailable for the remaining three tissues.

Evaluation of the 102 candidates using GNPC proteomic data showed that, among 37 proteins with available measurements, 21 showed significantly differential abundance after FDR correction (FDR < 0.05) in at least one stratum in plasma (Supplementary Table 10). In CSF, nine proteins showed nominally significant associations (p < 0.05), although none survived FDR correction, likely due to limited sample sizes (78 cases; Supplementary Table 11). Nine showed consistent associations in both sexes, including *SNCA* (target protein: α-synuclein), *NSF* (vesicle-fusing ATPase), and *CTSB* (cathepsin B). Female-predominant genes, such as *CTF1* (cardiotrophin-1) and *STX4* (syntaxin-4), also exhibited female-specific associations at protein level, with *CTF1* surviving FDR correction in females and *STX4* showing nominal significance (p = 0.031, FDR = 0.062). Additionally, *CD160*, a male-specific gene, was nominally elevated only in male cases (p = 0.045, FDR = 0.156), while another male-specific gene *L3MBTL2* (L3MBTL histone methyl-lysine binding protein 2) was elevated in both sexes.

Pathway enrichment analysis using the Reactome database identified distinct biological signatures by sex (Supplementary Table 12 and Supplementary Figure 3). Among all candidates significant from multiple analyses, WDR5-related chromatin regulation pathways were the most prominent signals surviving FDR correction in females, driven by *KANSL1, SETD1A,* and *KAT8*. At a nominal threshold (p < 0.01), additional female pathways included trans-Golgi network vesicle budding (driven by *STX4, HIP1R,* and *GAK*), chromatin modifying enzymes, and epigenetic regulation of gene expression. In males, SUMOylation-related pathways were identified at nominal significance, alongside Hurler syndrome-related pathways (driven by *IDUA* alone) in both sexes. Restricting the analysis to sex-specific candidates further reinforced these divergences: male signals were dominated by immune-related processes (FDR < 0.05), including pathogen recognition receptor signaling (driven by *CD160, FCGR2A,* etc.), while female signals were characterized by extracellular matrix organization, PI3K/AKT signaling, and interleukin-10 signaling in both sexes (Supplementary Figure 4 and Supplementary Table 13).

To further prioritize the most robust candidates, we identified eleven genes that reached significance across all three multi-omic analyses, either from sex-stratified or combined datasets. Of these, *SNCA, MAPT,* and *CTSB* were significant in both sexes, while *LRRC37A2* (mapping to the *MAPT* locus)*, GPNMB,* and *CD160* showed male-specific predominance, and *PRSS53* and *STX4* showed female-specific predominance. *BST1*, *SCARB2*, and *LGALS3* reached triple significance only in the sex-combined analysis. For these eleven candidates, we compared direction of effects across TWAS and PWMR (Figure 4), excluding gene-based association Z-scores, which reflect the strength rather than the direction of gene-trait association. For TWAS, where some genes showed tissue-dependent directionality, we reported the median Z-score from significant tissues. Notably, *SNCA* and *PRSS53* exhibited discordant effect directions between TWAS and PWMR, with positive associations in TWAS but negative (protective) associations in PWMR. In the case of *SNCA*, however, the whole blood TWAS result was concordant with the PWMR finding, both suggesting a protective effect, whereas predicted expression in other tissues was associated with increased PD risk, suggesting a tissue-dependent mechanism.

**Figure 4.**
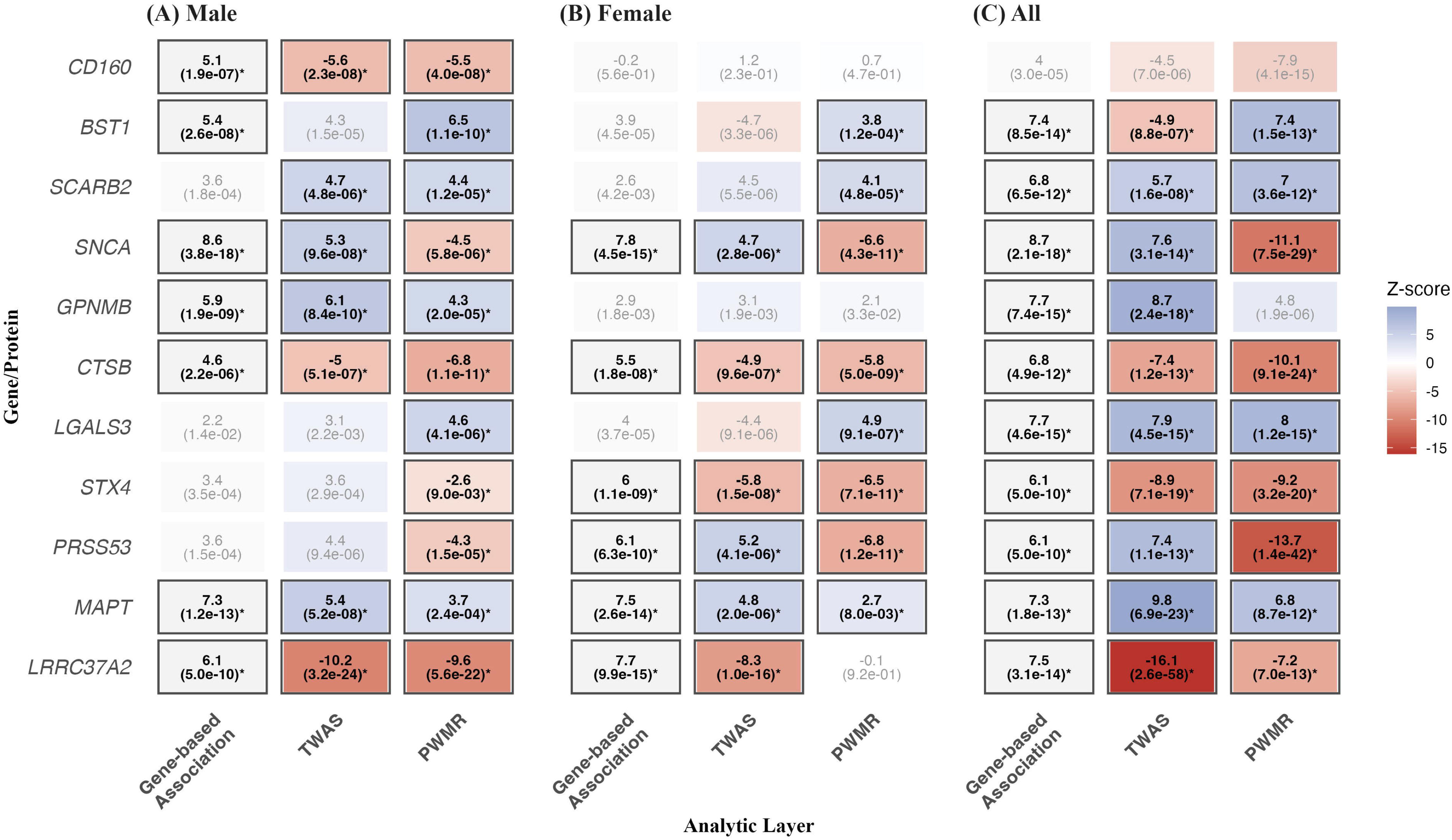
Multi-omic integration across genomic, transcriptomic, and proteomic layers. The heatmap illustrates standardized association strengths (Z-scores) across three distinct analytic layers: Gene-based association analysis, transcriptome-wide association study (TWAS), and proteome-wide Mendelian randomization (PWMR). Results are stratified into (A) male, (B) female, and (C) all PD cohorts to characterize sex-specific genetic architectures. Each cell displays the calculated Z-score with its corresponding p-value in parentheses. The gene-based association layer reports aggregate association strength without directional inference. For the TWAS and PWMR layers, Z-scores denote the direction of effect, where positive values (blue) indicate that increased gene expression or protein levels are associated with elevated PD risk, while negative values (red) indicate associations with decreased levels or protective effects. For the TWAS layer, the reported values represent median statistics derived from tissues achieving significance. Cells marked with asterisks (*) and highlighted borders indicate associations reaching statistical significance.

We visualized these eleven protein levels in plasma (Supplementary Figure 5) and CSF (Supplementary Figure 6). Several proteins exhibited opposite directions of association between the two biofluids. For *SNCA*, which showed discordant directions in TWAS (negative in whole blood and positive in other tissues) and PWMR (negative) analysis, plasma levels were higher in PD cases (OR = 1.44, p < 1×10^−30^), whereas CSF levels were lower in male PD (OR = 0.62, p = 0.007) with no association in females. *CTSB*, which showed negative associations in both TWAS and PWMR, exhibited higher plasma levels in PD cases but lower CSF levels only in females. *LGALS3* (galectin-3) was lower in the plasma of female PD patients, while *MAPT* (tau) was significantly elevated in the CSF of males and sex-combined group.

## Discussion

In this study, we conducted a comprehensive sex-stratified multi-omic integration — combining gene-based association, TWAS, and PWMR with colocalization, brain eQTL-based MR, and proteomic validation — to define the sex-specific molecular landscape of PD. Our analysis prioritized 102 candidates across 31 loci, of which eleven reached significance across all three analytical layers. Among these, *SNCA, MAPT,* and *CTSB* represented shared pathogenic axes across sexes, while clear sexual dimorphism emerged: *CD160* and *GPNMB* were male-predominant, implicating innate immune pathways, whereas *PRSS53* and *STX4* showed female predominance at the 16p11.2 locus. Pathway analysis further reinforced this divergence, revealing male-specific enrichment in SUMOylation and immune signaling versus female-specific WDR5-mediated chromatin remodeling. These findings demonstrate that PD is a molecularly heterogeneous disorder where the primary drivers differ between men and women.

Among the novel male-specific loci, *CD160* was supported by particularly robust convergent evidence across all analyses including colocalization and protein abundance analyses, with the sole exception of CSF protein abundance. CD160 is an activating receptor on natural killer (NK) cells that is essential for IFN-γ production.^31^ This finding is particularly relevant given recent evidence that NK cells play a pivotal role in clearing extracellular α-synuclein aggregates,^32^ and their depletion exacerbates synuclein pathology *in vivo*.^33^ In our TWAS and PWMR, genetically predicted lower CD160 levels were associated with increased PD risk in males, consistent with impaired NK cell-mediated protective clearance, while protein abundance analysis showed nominally increased levels in the plasma of male PD patients. The observed discrepancy — where genetic predisposition toward lower expression confers risk, yet actual protein levels are elevated — has been observed for other PD-associated proteins and may reflect reactive or compensatory changes in the disease state rather than causal biology. Although *CD160* has not been reported from genetic association studies of PD itself, it was identified in a genetic correlation study between PD and Crohn’s disease,^34^ supporting an immune-mediated link.

Another male-predominant candidate, *GPNMB* (glycoprotein non-metastatic melanoma protein B), emerged from all three analytical layers and has previously established associations from PD GWAS.^35^ *GPNMB* is a transmembrane glycoprotein highly expressed in microglia and macrophages, where it mediates phagocytic clearance and lysosomal function. *GPNMB* has been linked to *GBA1*-associated lysosomal dysfunction,^36^ the most common genetic risk factor for PD, and is secreted via *LRRK2*-modulated lysosomal exocytosis, with elevated CSF levels observed in both *LRRK2* and *GBA1* mutation carriers.^37^ The convergence of *CD160*, *FCGR2A*, and *GPNMB* within male-specific immune signaling enrichment further supports the predominant role of innate immune pathways in male PD pathogenesis.

*L3MBTL2* was prioritized as a novel male-specific risk gene, with evidence from gene-set and TWAS analyses. This gene encodes a component of the Polycomb repressive complex (PRC1.6), an epigenetic machinery that maintains gene silencing through chromatin compaction and histone modification.^38^ Dopamine signaling in parkinsonian models displaces Polycomb complexes (PRC1/2) from chromatin, leading to aberrant activation of genes associated with dyskinesia.^39^ Notably, WDR5, which also participates in the Polycomb complex, was the top female-enriched pathway signal in our study (driven by *KANSL1, SETD1A,* and *KAT8*), suggesting that epigenetic dysregulation through overlapping chromatin regulatory machinery may contribute to PD in both sexes through distinct molecular components. In addition, *L3MBTL2* has been identified as a pleiotropic locus shared between PD and depression,^40^ and among pleiotropic risk loci for neuroticism across eight psychiatric disorders.^41^ In our analysis, plasma protein levels were elevated in PD patients of both sexes, with a more pronounced increase in females.

In females, eight regions were prioritized from multiple analyses, with no novel loci identified, partly due to limited statistical power (12,054 male cases versus 7,384 female cases). However, *STX4* and *PRSS53* at the 16p11.2 locus reached significance across all three analytical layers in the female-stratified analysis, representing the most robust female-predominant candidates. The same locus also harbored *CTF1* and *KAT8*. Notably, *STX4* and *PRSS53* — along with several other PD-associated genes — have recently been implicated in autophagosome-lysosome fusion through tissue-specific isoform expression.^42^ Although their co-localization at 16p11.2 limits the ability to disentangle independent causal effects, our colocalization analysis provided partial resolution: *CTF1* and *STX4* shared a common causal variant in females (PP_H4_ > 0.9). *CTF1* (target protein: cardiotrophin-1), is a member of the IL-6 cytokine family that supports the survival of dopaminergic neurons,^43^ and *STX4* (syntaxin-4) mediates vesicle trafficking including dopamine transporter surface expression.^44^ These molecules showed opposite directions of effect between PWMR and protein abundance analysis: *CTF1* showed risk-increasing genetic associations but decreased protein levels in female PD, whereas *STX4* exhibited the reverse pattern. The female-dominant molecular landscape at this locus may be biologically related to estrogen-mediated neuroprotection in PD. *KAT8*, a histone acetyltransferase at the female-dominant 16p11.2 locus, participates in chromatin regulatory complexes that interact with estrogen receptor signaling.^45^ Supporting a potential hormonal link, our PWMR analysis identified *SHBG* (sex hormone-binding globulin) as protective against PD, with a more pronounced effect in females. *SHBG* is a key regulator of bioavailable sex hormones, and its protective association is consistent with the hypothesis that sex hormone signaling modulates PD risk in a sex-dependent manner. Nonetheless, the precise relationship between these genomic signals and estrogen-mediated neuroprotection cannot be resolved at the level of summary-level genetic analyses alone.

At a separate locus (14q22.3), *LGALS3* reached significance across all three analytical layers in the sex-combined analysis. Although *LGALS3* was not significant in sex-stratified analysis, plasma proteomic validation revealed a striking female-specific decrease in galectin 3 levels (OR = 0.61, p < 1 × 10^−30^) that was absent in males, suggesting a sexually dimorphic protein response. As a microglial-derived lectin implicated in α-synuclein aggregation and neuroinflammation,^46^ *LGALS3* may represent an underappreciated female-relevant target warranting further investigation.

For candidates that reached significance across multiple analytical layers, we further examined their protein levels in plasma and CSF data, alongside brain eQTL-based MR results, to assess the convergence of evidence across biological compartments. α-synuclein showed consistently elevated plasma levels in PD cases across both sexes, while CSF levels were nominally decreased in males (p = 0.007). This plasma–CSF discordance is consistent with the previous finding that CSF α-synuclein is already reduced at baseline in both manifest and prodromal PD,^47^ likely reflecting altered α-synuclein turnover — such as sequestration into aggregates — rather than ongoing dopaminergic neurodegeneration. Despite its robust associations in plasma pQTL-based PWMR, *SNCA* showed no significant brain eQTL-based MR associations. This may reflect the limited statistical power of brain eQTL datasets, restricted tissue coverage, or the composition of the MetaBrain samples, which include participants from neurodegenerative disease cohorts whose disease-related transcriptomic alterations may influence eQTL estimation. Tau protein showed no differential abundance in plasma but elevated CSF tau levels in male PD. However, the CSF data were derived from only two cohorts with limited sample sizes (41 male and 37 female cases), precluding reliable sex-stratified interpretation. It should be noted that the observed discordance between genetically predicted molecular effects (from TWAS and PWMR) and observed protein abundance in case-control cohorts (from GNPC data) likely reflects the fundamentally different causal frameworks of MR and cross-sectional analyses.

Our pathway enrichment analysis revealed distinct biological mechanisms by sex, with notable male-specific enrichment of SUMOylation. SUMOylation is a post-translational modification in which small ubiquitin-like modifier (SUMO) proteins are conjugated to target proteins, altering their stability, localization, or interactions. In the context of PD, SUMOylation of α-synuclein modulates its aggregation propensity and toxicity.^48^ Furthermore, *MAPT* (target protein: tau) is itself a target of SUMO modification, with SUMOylation implicated in tau stabilization and accumulation in neurofibrillary tangles.^49^ Male PD was also enriched in immune signaling, driven by multiple molecules including the previously described *CD160* and *FCGR2A*. *FCGR2A* encodes a low-affinity IgG Fc receptor on microglia whose causal variants map to microglia-specific enhancers, and also showed significant sexually dimorphic causal effect on PD from previous sex-stratified MR,^14^ with increased levels conferring risk specifically in males. In contrast, female PD was robustly enriched in WDR5-mediated chromatin remodeling. WDR5 is a core subunit of histone H3 lysine 4 (H3K4) methyltransferase complexes that regulate permissive gene expression in the brain, and its chromatin-binding activity is directly modulated by monoamine neurotransmitter-derived histone modification,^50^ suggesting a link between neurotransmitter signaling and epigenetic dysregulation in female PD.

Taken together, these findings support an integrative molecular model of sexually dimorphic PD pathophysiology. Male PD is characterized by convergent dysfunction across innate immune surveillance, post-translational modification via SUMOylation, and epigenetic repression. Female PD, by contrast, is characterized by chromatin remodeling and autophagosome-lysosome fusion. Despite these distinct molecular entry points, both sexes converge on shared pathogenic outcomes — lysosomal dysfunction (*CTSB*), vesicle trafficking disruption (*SNCA* and *MAPT*), and epigenetic dysregulation — consistent with a model of sex-modulated convergence in PD pathogenesis.

Several limitations should be acknowledged. First, the sex-stratified GWAS data from IPDGC has a smaller sample size, particularly for females, which may have reduced power to detect female-specific signals. Second, a common limitation shared by PWMR studies, including ours, is the use of sex-combined pQTL data as exposures, which may not fully capture sex-specific genetic regulation of protein levels. However, in our previous study using sex-stratified pQTL data,^15^ we observed results consistent with those obtained from sex-combined instruments, suggesting that sex-specific pQTL heterogeneity is unlikely to account for the observed sex-differential effects. Third, our analyses were restricted to individuals of European ancestry, limiting generalizability to other populations. Fourth, the proteomic validation relied on the SomaScan platform, which measures aptamer-based protein binding rather than absolute protein concentrations and may not distinguish between protein isoforms or post-translationally modified species, while our PWMR instrumental variables were derived from Olink-based pQTL data, introducing potential cross-platform discordance. Fifth, the CSF proteomic data were available from only two cohorts with limited sample sizes, and the GNPC constituent cohorts were not uniformly characterized in terms of disease duration, medication use, or clinical severity, precluding the distinction between causal and disease-driven protein changes.

In conclusion, our integrative multi-omic analysis reveals that PD is a sexually dimorphic disorder at the molecular level. Male PD is characterized by immune dysregulation alongside SUMOylation, with *CD160* and *L3MBTL2* emerging as novel male-specific candidates. Female PD is associated with chromatin remodeling, with female dominance observed in 16p11.2 and 14q22.3 loci. These findings underscore the necessity of sex-stratified approaches in biomarker discovery and therapeutic development for PD.

## Acknowledgements

We acknowledge the participants and investigators of the IPDGC, GP2, UKB-PPP, GTEx, MetaBrain and GNPC consortia for generating and making their data publicly available.

## Supplementary material

Supplementary material is available at *Brain* online.

